# PI3K/mTOR and topoisomerase inhibitors with potential activity against SARS-CoV-2 infection

**DOI:** 10.1101/2020.09.02.20186783

**Authors:** James Robert White, Michael Bonner Foote, Justin Jee, Guillem Argilés, Jonathan C.M. Wan, Luis Alberto Diaz

**Affiliations:** Resphera Biosciences, Baltimore, MD USA; Department of Medicine, Memorial Sloan Kettering Cancer Center, New York, NY, USA

## Abstract

There is an urgent need to identify therapies to prevent and treat SARS-CoV-2 infection. We performed a statistical evaluation of in vitro gene expression profiles reflecting exposure to 1,835 drugs, and found topoisomerase inhibitors and PI3K/mTOR pathway inhibitors among the strongest candidates for reduced expression of ACE2, a host gene associated with SARS-CoV-2 infection. Retrospective clinical data suggest that patients on these agents may be less likely to test positive for SARS-CoV-2.

## Introduction

Severe acute respiratory syndrome coronavirus 2 (SARS-CoV-2) continues to spread globally with over 13.1 million diagnosed cases and 574,000 deaths (as of 14 July, 2020). SARS-CoV-2 is a single-stranded RNA virus that utilizes spike (S) glycoproteins to infect individuals by attaching to angiotensin-converting enzyme 2 (encoded by the ACE2 gene) expressed on airway epithelia^1–3^. After attachment, hemagglutinin cleavage of ACE2 initiates viral internalization and subsequent viral S protein cleavage by the transmembrane protease serine 2 TMPRSS2 to enable viral entry^3^. Other transmembrane proteases, such as ADAM17 and TMPRSS11D, may also influence cleavage of ACE2^4,5^.

ACE2 gene expression may provide a unique therapeutic target to help prevent SARS-CoV-2 infection. Hofman et al. found that ACE2 expression on a panel of cell lines correlated with *in vitro* susceptibility to SARS-CoV S-driven infection^4^. Many ACE2-associated factors, including TMPRSS2, have no recognized indispensable somatic functions; drugs targeting this pathway may show a promising therapeutic ratio^6^.

The Library of Integrated Network-Based Cellular Signatures (LINCS) program is an open resource for the scientific community that includes expression profiles of cells exposed to a variety of perturbing agents^7^ including small molecules over a range of concentrations. Here, we report on a comprehensive statistical evaluation of cell line gene expression signatures associated with various drug exposures in LINCS. We analyzed 1,835 drugs for which distinct cells lines and variable dosages were available, and sought to identify those with statistically significant reductions in expression of genes associated with SARS-CoV-2 infection. We then utilized retrospective clinical data to evaluate whether cancer patients receiving drugs associated with reduced expression of ACE2 demonstrated a decreased odds ratio for a positive SARS-CoV-2 test compared to patients receiving other antineoplastic therapies.

## Results and Discussion

For each drug, we fitted a generalized linear model to moderated Z-score values^8^ reflecting relative ACE2 gene expression across seven cell lines and drug dosages from 0.01 uM to 10 uM (Table S1). Among those drugs with the strongest associated reductions in ACE2 expression, we identified two major categories: (i) topoisomerase inhibitors, and (ii) PI3K/mTOR pathway inhibitors (Table 1). Topoisomerase inhibitors including camptothecin, SN-38, and Genz-644282 demonstrated significant reductions in ACE2 expression, often with a dose dependent relationship (Fig. 1). PI3K/mTOR inhibitors including PF-04691502, GDC-0980(RG7422), and Taselisib also displayed dose dependent reductions in ACE2 expression (Fig. 2). Moreover, two AKT inhibitors related to the PI3K/mTOR pathway, Afuresertib and MK-2206, were associated with reduced ACE2 (Table 1).

**Figure 1:**
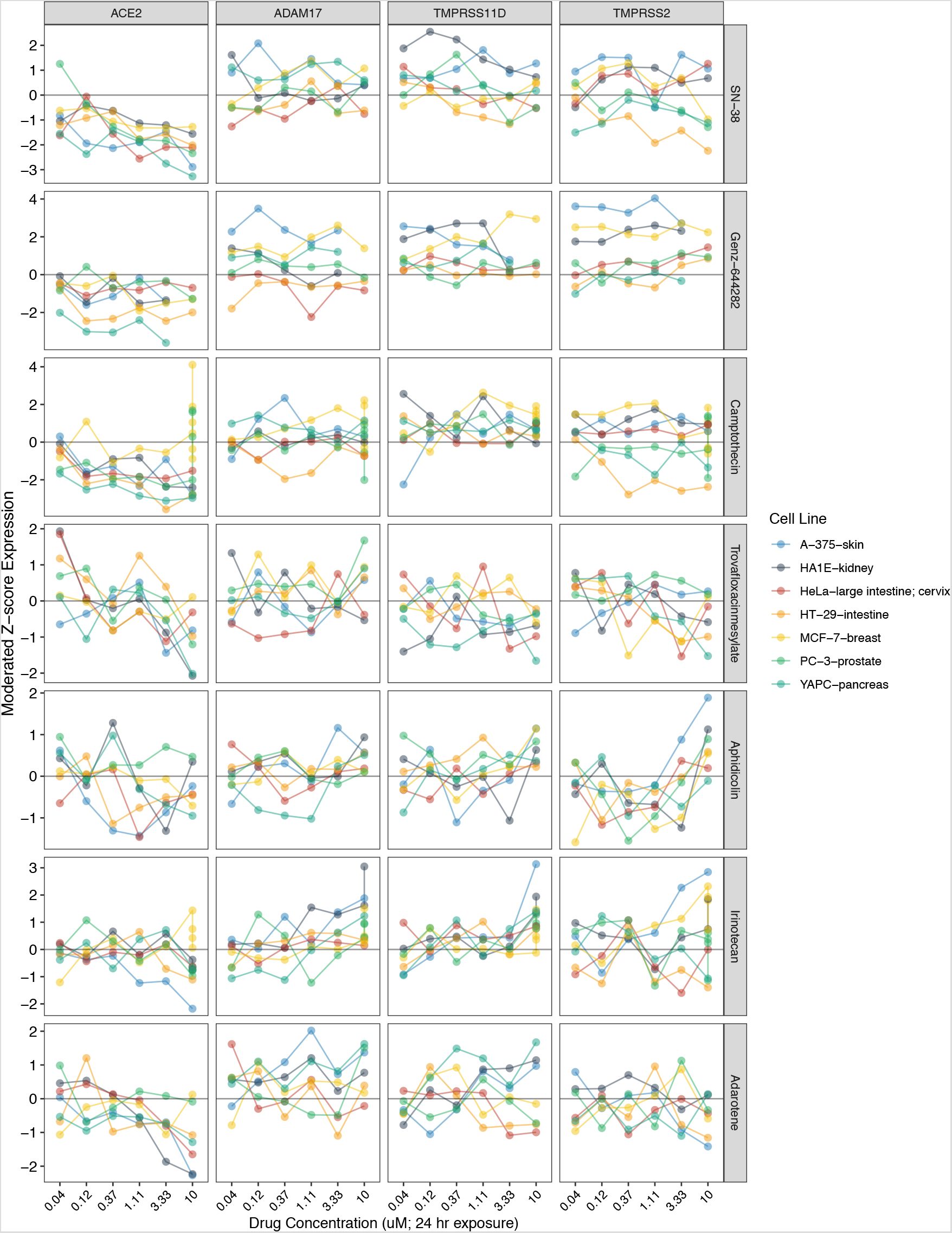
ACE2 expression reductions associated with exposure to various topoisomerase inhibitors. Related genes (ADAM17, TMPRSS11D, TMPRSS2) included for comparison. Y-axis (left) = moderated Z-scores summarizing differential expression across multiple replicates per cell line calculated by the LINCS project. Y-axis (right) = drug name.

**Figure 2:**
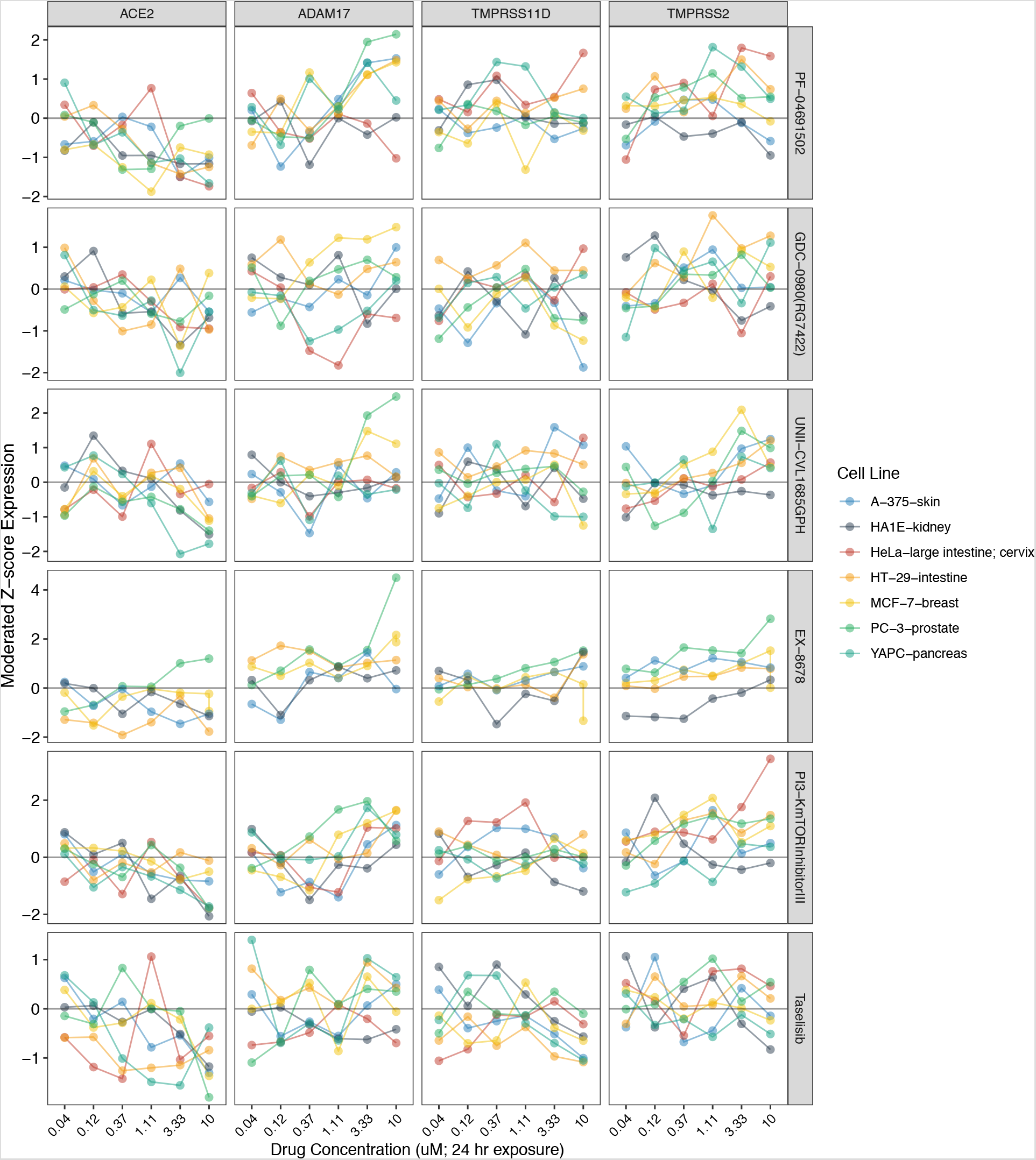
ACE2 expression reductions associated with exposure to PI3K/mTOR inhibitors. Related genes (ADAM17, TMPRSS11D, TMPRSS2) included for comparison. Y-axis = moderated Z-scores summarizing differential expression across multiple replicates per cell line calculated by the LINCS project. Y-axis (right) = drug name.

**Table 1.**
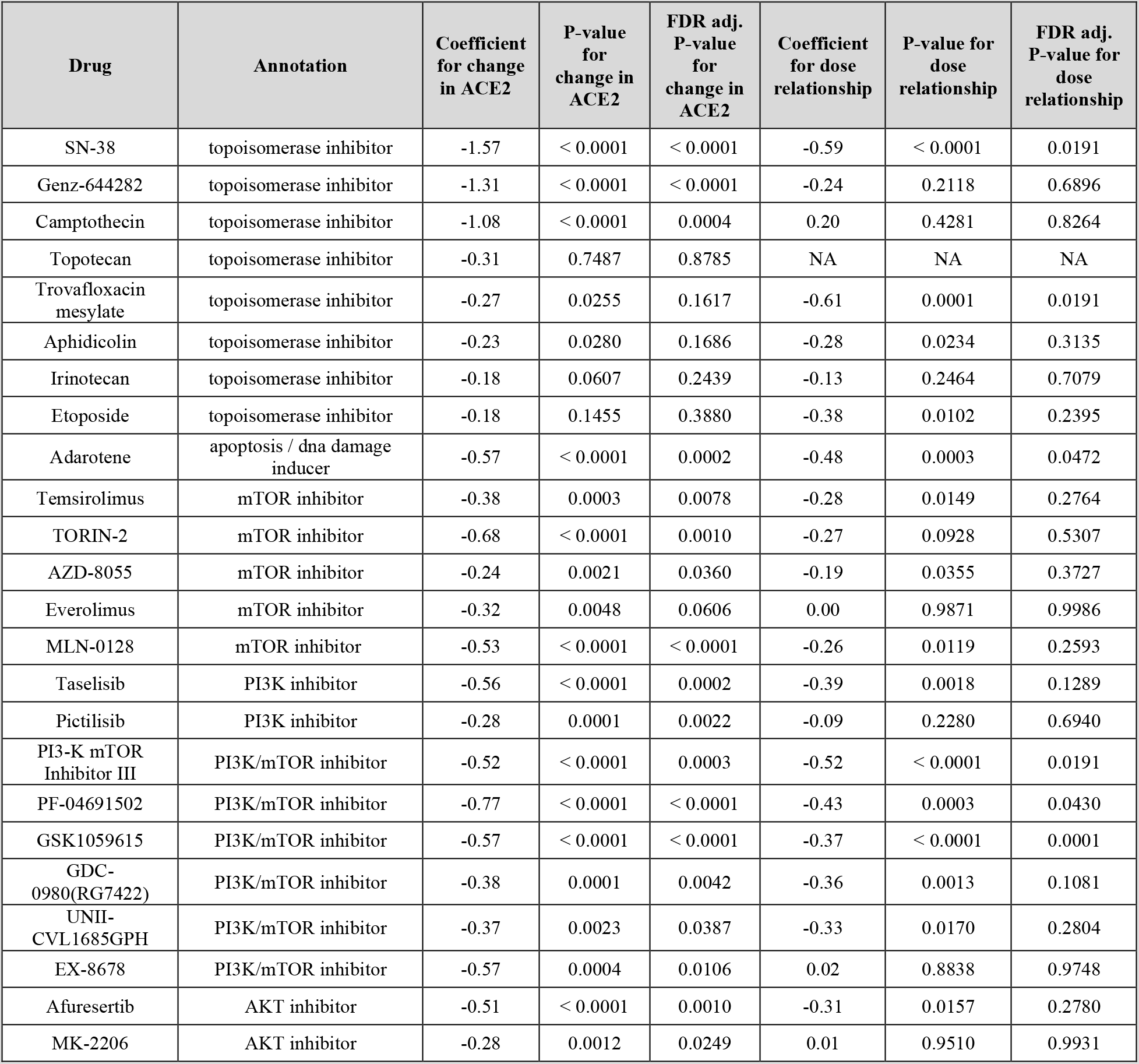
Classes of drugs commonly identified as reducers of ACE2 expression. Generalized fixed effects model results reported per drug including coefficient for ACE2 expression (baseline) and associated with drug dose relationship. “N/A” designates “Not Applicable”. Unadjusted and false discovery rate (FDR) adjusted P-values are included.

We next performed a retrospective review of clinical records to evaluate the frequency of SARS-CoV-2 infection in patients on these ACE2-associated antineoplastics. Retrospective data was obtained from an IRB-approved study of adult cancer patients tested for SARS-CoV-2 receiving active antineoplastic therapy at Memorial Sloan Kettering Cancer Center (MSKCC) during the COVID-19 epidemic period (n = 4,040 patients; Table S2). The overall study population had mainly solid tumors (n = 3771; 93.3%). Thirteen percent (n = 535) of the study population was actively treated with an ACE2-associated drug (Table S2, Table S5). Patients treated with ACE2-associated drugs showed similar clinical demographics to those treated with different antineoplastics (Table S2).

Patients receiving ACE2-associated therapies demonstrated a lower univariate odds ratio (0.65, 95% CI 0.00–0.98, P = 0.04) for a positive SARS-CoV-2 test during active antineoplastic therapy compared to patients on other agents (Table S3). Additional study covariates shown to be significant in cancer patients were also evaluated. Patients with non-white race (OR 2.00), hematologic malignancy (OR 2.89) and metastatic disease (OR 1.49) also demonstrated statistically significant increase in the odds of a positive SARS-CoV-2 test (Table S3). Significant covariates were placed into a multivariate logistic regression model (Table S4). Active therapy trended towards independent significance for a SARS-CoV-2 positive test (OR 0.68, 95% CI 0.40–1.07, BH-corrected q = 0.13).

Our results reveal several antineoplastic agents that may demonstrate reductions in ACE2 expression, a potential target proposed in preventing and treating SARS-CoV-2 infection. Cancer patients taking these potential ACE2-associated agents showed lower rates of a positive SARS-CoV-2 test compared to patients taking other forms of active antineoplastic therapy. These medications were universally held when patients tested positive for SARS-CoV-2, and our study was underpowered to detect possible differences in COVID-19 associated endpoints, such as admission, hypoxic events and death.

These findings are consistent with recent studies that propose topoisomerase inhibitors or PI3K/mTOR inhibitors for treatment of COVID-19. One recent study proposed Irinotecan and Etoposide combination therapy for critically ill COVID-19 patients^9^ based on the immunomodulatory and viral suppressive profiles of these drugs. Additionally, a recent loss-of-function experiment revealed that certain topoisomerases were required for efficient replication of positive-sense RNA viruses including SARS-CoV-2^10^. Immunoregulatory and anti-viral properties have also been cited as rationales for PI3K/mTOR pathway inhibitors to treat COVID-19^11–17^.

This study has several limitations. ACE2-associated antineoplastic therapies were discovered by *in-vitro* computer modeling and need to be validated experimentally. Although *in vitro* data implicates ACE2 in the SARS-CoV-2 infection route, ACE2 is not yet a clinically validated target. Furthermore, clinical data obtained were retrospective and may be underpowered to estimate the true impact of ACE2-associated therapy on SARS-CoV-2 positivity and COVID-19 adverse outcomes. Finally, study patient characteristics are skewed towards non-metastatic (63.2%), solid tumor (93.3%) patients. Despite these limitations, our results suggest that further investigation of PI3K/mTOR pathway inhibitors and topoisomerase inhibitors as a COVID-19 preventative strategy are warranted.

## Methods

### In Vitro Modeling and Analysis

LINCS gene expression signatures and associated drug exposure metadata were collected using the LINCS Data Portal API (http://lincsportal.ccs.miami.edu/apis). Compounds considered were limited to those tested on the seven specific cell lines of interest with 24 hours of exposure time at concentrations ranging from 0.01uM to 10uM. For each candidate drug, a generalized linear model was fit to ACE2 moderated Z-scores values adjusted for log10(drug concentration). Calculated by LINCS, moderated Z-scores reflect a weighted average of replicate samples in an experiment of a specific cell line and drug concentration. Model coefficients and P-values associated with baseline expression and drug concentration (two-sided test; null hypothesis *b* = 0) were used to prioritize drugs with significant reductions in ACE2 expression and dose dependent relationships. P-values were adjusted for multiple hypothesis testing using the false discovery rate (FDR).

### Data Availability

LINCS Phase I data are publicly available through NCBI (GEO accession GSE92742) and the LINCS Data Portal 2.0 (http://lincsportal.ccs.miami.edu/). Procedures on LINCS signature generation and moderated Z-score calculation can be found in the GEO LINCS user guide (v2.1; https://docs.google.com/document/d/1q2gciWRhVCAAnlvF2iRLuJ7whrGP6QjpsCMq1yWz7dU/edit#). Aggregated patient data used in this study is available upon request from the corresponding author (L.A.D.). Requests will be first reviewed for compliance with the ethical and patient privacy regulations of the Memorial Sloan Kettering Cancer Center.

### Code Availability

The R code used for this study is available upon request from the corresponding author.

### Clinical Data Acquisition and Definitions

Electronic medical record (EMR) auto-populated flowsheet data was obtained for adult cancer patients at Memorial Sloan Kettering Cancer Center (MSKCC) undergoing active antineoplastic treatment during the time of the first positive SARS-CoV-2 tests (03/01/2020) with a study data freeze on 5/28/20 for analysis. Patients with antineoplastic therapies that were discontinued before this study period were not included in the study. Auto-populated flowsheet data was obtained from a standardized-input curated database that assessed the time and date of antineoplastic administration, as well as patient demographics, disease characteristics and study endpoints. SARS-CoV-2 testing was performed at MSKCC using a nasopharyngeal swab to determine the presence of virus specific RNA (MSKCC FDA-EUA-approved assay, and commercial assay Cephied®). Patients who had a positive test were classified as SARS-CoV-2 positive. Patients who tested negative or were not tested for SARSCoV-2 were classified as “Not SARS-CoV-2 positive”.

Study subgroups were classified as in Table S2. Notably, malignancy status as solid or hematologic was not mutually exclusive; several study patients had both an active solid and liquid malignancy, and were thereby coded as having each separate subtype. Although most patients were coded for the study variables, several patients did not have coded demographics and these patients were not associated with their respective missing demographic. For example, 89.1% of patients were coded for tobacco status with 10.9% of patients with an unknown tobacco status; these patients were not associated with a tobacco status in the statistical analysis.

Antineoplastic therapies were grouped using standardized antineoplastic drug categories (Table S5). Antineoplastic agents with significant activity from *in vitro* assessment (Table 1) were cross-referenced against the study population to determine ACE2-associated therapy status. If a patient received an ACE2-expression reducing therapy during the study period (03/01/20–05/28/20), they were classified in the “ACE2-associated antineoplastic” subgroup. Patients who did not receive a potential ACE2 expression reducing agent were classified in the “No ACE2-associated antineoplastic” subgroup.

### Clinical Data Statistical Analysis

Univariate, one-sided Fisher’s exact statistical testing was used to test the specific hypothesis that active ACE2-associated antineoplastic use is associated with a lower odds ratio of a positive SARS-CoV-2 test. Additional study covariates (female gender, age greater than or equal to 65, non-white race, smoking, hematologic malignancy, metastatic disease) were similarly evaluated. Odds ratios, 95% confidence intervals, and one-sided p-values were reported for each covariate. Covariates with a p-value of less than or equal to 0.10 were placed into a multivariate model, and logistic regression was performed. To account for multiplicity, p-values for each covariate in the multivariate model were subject to Benjamini-Hochberg correction and q-values were reported.

The odds ratios of achieving specific clinical endpoints, including admission, hypoxic event (defined as requiring greater than 4 L of oxygen), and death, were assessed comparing patients with ACE2-associated antineoplastics and patients who did not received ACE2-associated antineoplastics using similar Fisher’s exact statistical testing. No significant difference in adverse COVID-19 outcomes (admission, hypoxia requiring greater than 4 liters of supplementary oxygen, death, or composite of these three outcomes) was observed in the overall study population, or a subset population of only SARS-CoV-2 positive patients.

All statistical computation was done with use of R Studio (version 1.2.5033) and R (v4.0.0) statistical software.

### Study Approval

This study was approved by the Institutional Review Board at Memorial Sloan Kettering Cancer Center.

## Data Availability

LINCS Phase I data are publicly available through NCBI (GEO accession GSE92742) and the LINCS Data Portal 2.0 (http://lincsportal.ccs.miami.edu/). Procedures on LINCS signature generation and moderated Z-score calculation can be found in the GEO LINCS user guide (v2.1; https://docs.google.com/document/d/1q2gciWRhVCAAnlvF2iRLuJ7whrGP6QjpsCMq1yWz7dU/edit#).Aggregated patient data used in this study is available upon request from the corresponding author (L.A.D.). Requests will be first reviewed for compliance with the ethical and patient privacy regulations of the Memorial Sloan Kettering Cancer Center. 

https://docs.google.com/document/d/1q2gciWRhVCAAnlvF2iRLuJ7whrGP6QjpsCMq1yWz7dU/edit

https://www.ncbi.nlm.nih.gov/geo/

## Author contributions

J.R.W. and L.A.D. conceived and designed the study. J.R.W. and M.B.F. analyzed and interpreted data. J.J. and M.B.F. collected retrospective patient data. J.C.M.W. and A.G. provided guidance and scientific input. J.C.M.W. and A.G. interpreted results. J.R.W., J.C.M.W., and M.B.F. drafted the manuscript. All authors read and approved the final manuscript.

## Acknowledgements

Research supported by a Stand Up to Cancer Colorectal Cancer Dream Team Translation Cancer Research Grant (SU2C-AACR-DT22–17). Stand up to Cancer is a program of the Entertainment Industry Foundation administered by the American Association for Cancer Research. M.B.F. is partially supported by a T32 NIH Scholar Grant.

---

**Supp Table. S1.** Differential expression results for ACE2 (LINCS Phase I data) using moderated Z-score signature measures of relative expression *[See supplementary Excel file]*

**Supp Table. S2:** Summary of overall study patient characteristicsDescriptions of variables in the meta-population model. Demographic, clinical and treatment characteristics of all study patients, stratified into subgroups by ACE2-associated antineoplastic therapy status are reported. *ACE2-associated therapies and other anti-neoplastic therapies are not mutually exclusive; patients within the subgroup could be exposed to multiple agents during the study period. “N/A” designates “Not Applicable”.

|  | All (n=4040) | No ACE2-associated antineoplastic(n=3505) | ACE2-associated antineoplastic (n=535) |
| --- | --- | --- | --- |
|  | No. (% Total Patients) | No. (% Column) | No. (% Column) |
| **Demographics** |  |  |  |
| Female | 2406 (59.6%) | 2095 (59.8%) | 311 (58.1%) |
| Male | 1634 (40.4%) | 1410 (40.2%) | 224 (41.9%) |
| White | 3179 (78.7%) | 2753 (78.6%) | 426 (79.6%) |
| Non-White | 703 (17.4%) | 613 (17.5%) | 90 (16.8%) |
| Age ≥ 65 | 2034 (50.3%) | 1786 (51.0%) | 248 (46.4%) |
| Age < 65 | 2006 (49.7%) | 1719 (49.1%) | 287 (53.6%) |
| Tobacco former or current smoker | 2034 (50.3%) | 1786 (51.0%) | 248 (46.4%) |
| Never smoked tobacco | 2006 (49.7%) | 1719 (49.1%) | 287 (53.6%) |
| **Cancer Overview** |  |  |  |
| Solid malignancy* | 3771 (93.3%) | 3273 (93.4%) | 498 (93.1%) |
| Hematologic malignancy* | 269 (6.7%) | 232 (6.6%) | 37 (6.9%) |
| Metastatic disease | 1485 (36.8%) | 1305 (37.2%) | 180 (33.6%) |
| Non-metastatic disease | 2555 (63.2%) | 2200 (62.8%) | 355 (66.4%) |
| **Outcomes** |  |  |  |
| SARS-CoV-2 positive | 207 (5.1%) | 188 (5.4%) | 19 (3.6%) |
| Not SARS-CoV-2 positive | 3833 (94.9%) | 3317 (94.7%) | 516 (96.4%) |
| Admission | 559 (13.8%) | 452 (12.9%) | 107 (20.0%) |
| Hypoxic event | 69 (1.7%) | 52 (1.5%) | 17 (3.2%) |
| Death | 133 (3.3%) | 110 (3.1%) | 23 (4.3%) |
| **ACE2-associated therapy\*** |  |  |  |
| Topotecan | 18 (0.4%) | N/A | 18 (3.4%) |
| Irinotecan | 281 (7.0%) | N/A | 281 (52.5%) |
| Etoposide | 74 (1.8%) | N/A | 74 (13.8%) |
| Temsirolimus | 6 (0.1%) | N/A | 6 (1.1%) |
| Everolimus | 151 (3.7%) | N/A | 151 (28.2%) |
| Alpelisib | 59 (1.5%) | N/A | 59 (11.0%) |
| Duvelisib | 7 (0.2%) | N/A | 7 (1.3%) |
| Idelalisib | 2 (0.1%) | N/A | 2 (0.4%) |
| MLN-0128 | 1 (0.1%) | N/A | 1 (0.2%) |
| Ipasertib | 1 (0.1%) | N/A | 1 (0.2%) |
| AZD5363 | 3 (0.1%) | N/A | 3 (0.6%) |
| **Other anti-neoplastic therapy \*** |  |  |  |
| Alkylating agents | 601 (12.3%) | 601 (17.1%) | N/A |
| Anti-CTLA-4 monoclonal antibodies | 55 (1.1%) | 55 (1.6%) | N/A |
| Anti-PD-1 monoclonal antibodies | 654 (13.4%) | 654 (18.6%) | N/A |
| Antimetabolites | 802 (16.4%) | 802 (22.9%) | N/A |
| Antineoplastic antibiotics | 186 (3.8%) | 186 (5.3%) | N/A |
| Multidrug formulations | 15 (0.3%) | 15 (0.4%) | N/A |
| BCR-ABL tyrosine kinase inhibitors | 65 (1.3%) | 65 (1.9%) | N/A |
| BTK inhibitors | 34 (0.7%) | 34 (1.0%) | N/A |
| CD20 monoclonal antibodies | 19 (0.4%) | 19 (0.5%) | N/A |
| CD30 monoclonal antibodies | 11 (0.2%) | 11 (0.3%) | N/A |
| CD38 monoclonal antibodies | 38 (0.8%) | 38 (1.1%) | N/A |
| CDK 4/6 inhibitors | 291 (6.0%) | 291 (8.3%) | N/A |
| EGFR inhibitors | 195 (4.0%) | 195 (5.6%) | N/A |
| Hedgehog pathway inhibitors | 2 (0.1%) | 2 (0.1%) | N/A |
| HER2 inhibitors | 223 (4.6%) | 223 (6.4%) | N/A |
| Histone deacetylase inhibitors | 2 (0.1%) | 2 (0.1%) | N/A |
| Mitotic inhibitors | 523 (10.7%) | 523 (14.9%) | N/A |
| Multikinase inhibitors | 388 (8.0%) | 388 (11.1%) | N/A |
| Other antineoplastic | 559 (13.8%) | 559 (16.0%) | N/A |
| Oral immunomodulatory | 107 (2.2%) | 107 (3.0%) | N/A |
| PARP inhibitors | 133 (2.7%) | 133 (3.8%) | N/A |
| Proteasome inhibitors | 47 (1.0%) | 47 (1.3%) | N/A |
| Topical chemotherapy | 17 (0.3%) | 17 (0.5%) | N/A |
| VEGF/VEGFR inhibitors | 330 (6.8%) | 330 (9.4%) | N/A |

**Supp Table. S3:** Univariate analysis of patients receiving active antineoplastic therapy establishes odds ratios for a SARS-CoV-2 positive test for study covariates. Univariate analysis of study covariates demonstrates a significant decreased odds ratio for a SARS-CoV-2 positive test in patients with receiving ACE2-associated antineoplastic therapy (OR 0.65, 95% CI 0.00–0.98, p-value 0.04).

| Covariate | OR | 95%<br>CI<br>Min | 95%<br>CI<br>Max | p-value |
| --- | --- | --- | --- | --- |
| ACE2-associated antineoplastic therapy | 0.65 | 0.00 | 0.98 | 0.04 |
| Female gender | 0.79 | 0.00 | 1.01 | 0.06 |
| Age greater than or equal to 65 | 1.04 | 0.81 | Inf | 0.43 |
| Non-white race | 2.00 | 1.51 | Inf | 0.00 |
| Smoking | 0.91 | 0.69 | Inf | 0.76 |
| Hematologic malignancy | 2.89 | 2.02 | Inf | 0.00 |
| Metastatic disease | 1.49 | 1.16 | Inf | 0.00 |

**Supp Table. S4:** Multivariate logistic regression of study covariates in patients receiving active antineoplastic therapy. Multivariate logistic regression analysis of covariates demonstrating statistical significance (p≤0.10) in univariate analysis were included in a multivariate logistic regression. ACE2-associated antineoplastic therapy shows a trend towards significance (OR 0.68, 95% CI 0.41–1.07, BH-corrected q-value of 0.13).

| Covariate | OR | 95%<br>CI<br>Min | 95%<br>CI<br>Max | p-value | BH-corrected<br>q-values |
| --- | --- | --- | --- | --- | --- |
| ACE2-associated antineoplastic therapy | 0.68 | 0.40 | 1.07 | 0.11 | 0.13 |
| Non-white race | 2.02 | 1.46 | 2.77 | 0.00 | 0.00 |
| Hematologic malignancy | 2.62 | 1.70 | 3.93 | 0.00 | 0.00 |
| Female gender | 0.76 | 0.57 | 1.03 | 0.07 | 0.12 |
| Metastatic disease | 1.26 | 0.93 | 1.69 | 0.13 | 0.13 |

**Supp Tab. S5:** Major antineoplastic subcategorization key. Patient classification mapping shown for specific antineoplastic drugs used by patients in the “No ACE2-associated antineoplastic cohort” and their respective antineoplastic subcategory used in Supplementary Table S2 *[See supplementary Excel file]*

